# Partial loss of *FITM2* function causes hereditary spastic paraplegia

**DOI:** 10.1101/2025.01.23.24319660

**Authors:** Ainara Salazar-Villacorta, Laura M. Bond, Leehyeon Kim, Katherine Anagnostopoulou, Annarita Scardamaglia, Eugenia Filippakopoulou, Athina Ververi, Stephanie Efthymiou, Argirios Dinopoulos, David Murphy, Georgia Karadima, Georgios Koutsis, Marios Kaliakatsos, Henry Houlden, Tobias C. Walther, Robert V. Farese

## Abstract

*FITM2* encodes fat-storage inducing transmembrane protein 2 (FIT2), a lipid diphosphatase in the ER that cleaves acyl-CoAs and is crucial for ER homeostasis. In humans, homozygous null mutations in *FITM2* are associated with a syndrome characterized by deafness and dystonia. Here, we report two families with hereditary spastic paraplegia (HSP) in whom exome sequencing revealed compound heterozygosity for *FITM2* mutations. In each family, the affected probands carry one putative null allele and one G100R missense allele. Functional analyses demonstrated that the G100R allele is hypomorphic, with FIT2 protein levels reduced to 20% of wild type, leading to proportionately decreased enzyme activity. The occurrence of similar HSP disease phenotypes and the same hypomorphic mutation in these families suggests that the G100R mutation and its associated reduced enzyme activity represent a newly recognized clinical manifestation of *FITM2* mutations, expanding the spectrum of conditions associated with this gene.

## INTRODUCTION

Since the first genetic loci associated with hereditary spastic paraplegia (HSP) were discovered in the 1990s, more than 80 genes causing HSP have been discovered.^1^ HSPs are a diverse group of neurological disorders with the common feature of bilateral lower limb spasticity and weakness as the predominant clinical manifestation, due to a length-dependent axonopathy of the upper corticospinal motor neurons.^2–4^

HSPs are categorized as pure or complicated. Pure HSP involves isolated spastic paraplegia of the lower limbs, urinary dysfunction, and reduced vibration sensation. Complicated HSP includes additional neurological and extra-neurological symptoms, such as epilepsy, cerebellar ataxia, cognitive impairment, and visual disturbances.^1–4^

Childhood-onset HSP often presents in complicated manners^5,6^, and its genetic landscape is complex due to the recent association of non-classical HSP genes with HSP phenotypes^7,8^, the rapid rate of gene discovery^8,9^, and the significant overlap with other neurological disorders, such as cerebellar ataxia, peripheral neuropathy, parkinsonism, and amyotrophic lateral sclerosis (ALS)^10^. As a result, over 150 different genes are now recognized as potential causes of childhood-onset HSP phenotypes on genomic platforms used for genetic diagnosis in clinical settings, such as the Panel App (“R61 Childhood onset hereditary spastic paraplegia”, Genomics England, 2024)^5^.

Many HSP genes encode proteins involved in membrane and organelle biology, and around 30 HSP genes are involved in lipid metabolism, making this one of the most common pathways affected in this group of disorders^2^.

Here, we report patients from two families with a complex HSP phenotype, characterized by predominant spasticity and associated cerebellar signs and caused by *FITM2* biallelic variants. *FITM2* encodes fat-storage-inducing transmembrane protein 2 (FIT2), a lipid diphosphatase thought to cleave acyl-CoAs within the ER membrane^11^. FIT2 activity is crucial for ER lipid homeostasis in cells and organisms^12,13^. Cells lacking FIT2 exhibit abnormal lipid droplet formation^14–18^, defects in lipid metabolism^19^, and ER stress^13,20^. The defects in LD formation most likely result as an indirect consequence of compromised catalytic activity^11,15,16,21,22^. Deleting the FIT2-encoding gene in species, such as *Caenorhabditis elegans* or *Mus musculus,* is lethal^15,23^. Selectively deleting *Fitm2* in hepatocytes of mice results in a pleiotropic phenotype of liver injury with ER stress and dysfunction^12,13^. In humans, complete deficiency of *FITM2* leads to a rare disease with deafness and dystonia called Siddiqi syndrome^24–27^.

Here, we determined the molecular etiology of HSP in two families. We found that they share *FITM2* mutations of compound heterozygosity, each with one *FITM2* null allele and the same missense variant that leads to reduced FIT2 abundance and activity. Thus, our results describe a new clinical cause of HSP due to mutations in the *FITM2* locus.

## RESULTS

### Clinical findings and case descriptions

Clinical observations from the two identified families (**Figure 1**) exhibiting the novel HSP complex phenotype are presented in **Table 1**, accompanied by a review of prior cases associated with *FITM2* variants and Siddiqi syndrome.

**Figure 1.**
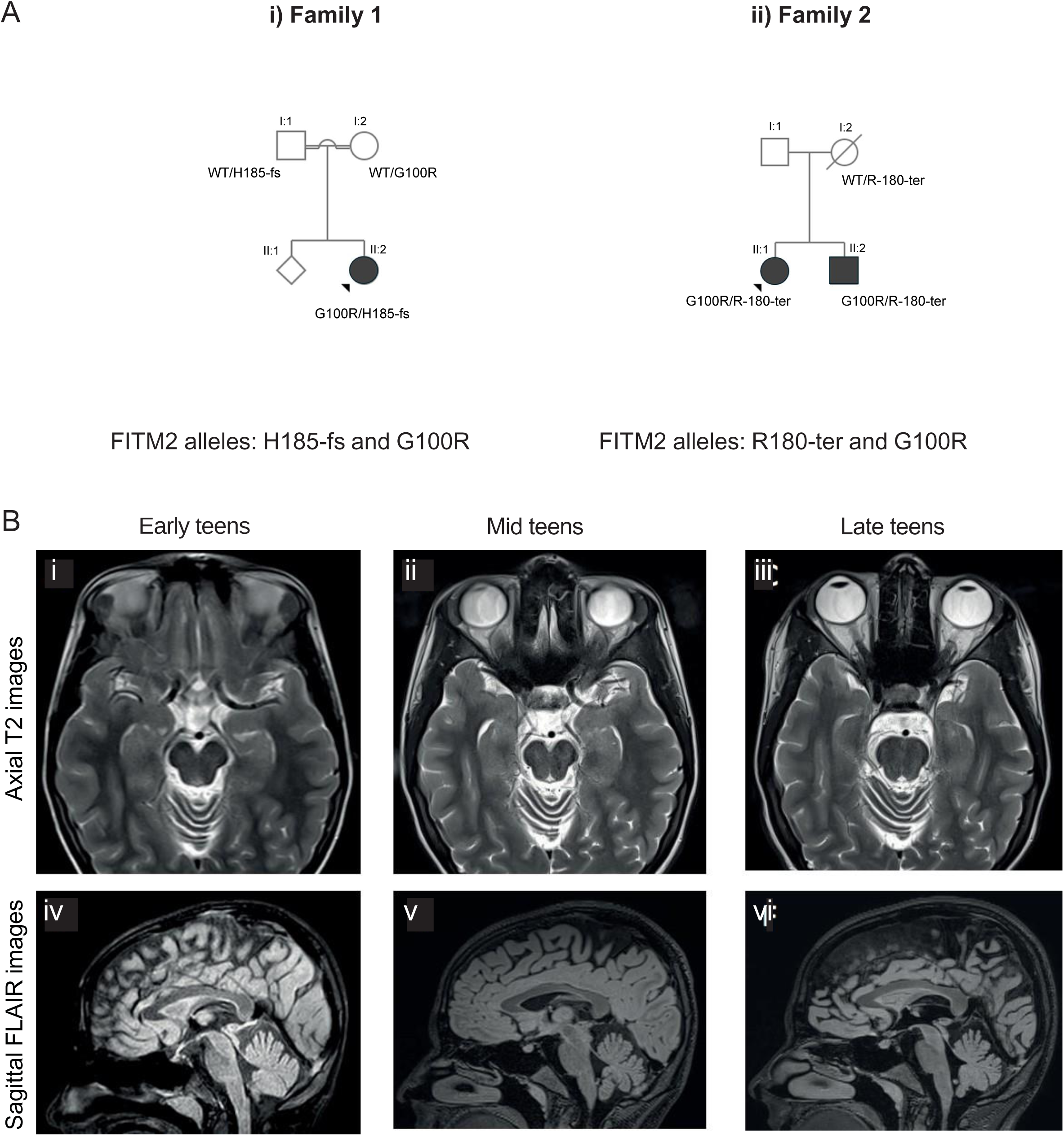
New FITM2 mutations reported. **(A)** Family pedigree of the newly reported families and variant segregation (i) Family 1 and (ii) Family 2. **(B)** Brain MRI characteristics. Sequential brain MRI images of the proband of Family 1 (II:2). (i-iii): Axial T2 images showing mild atrophy of the cerebellar vermis. (iv-vi): Sagittal FLAIR images showing mild atrophy of the cerebellar vermis. There is minimal change over the years (i and iv, early teens; ii and v, mid teens; iii and vi: late teens).

**Table 1.**
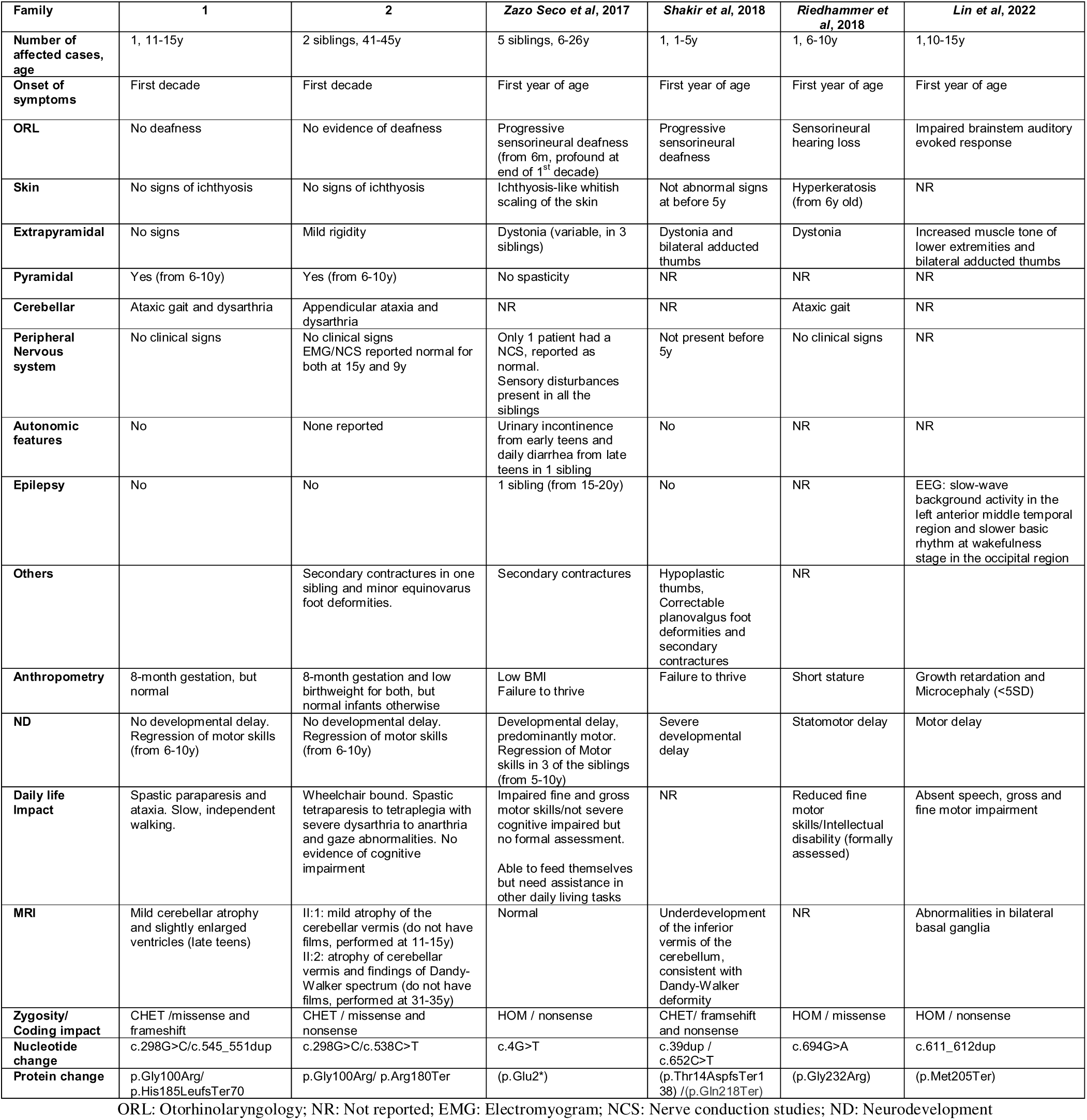
Clinical features and *FITM2* variant characteristics in families 1 and 2, with a review of families reported in the literature.

#### Family 1

The proband and index case (II:2, Family 1) was a female aged between 16 and 20. There was consanguinity in the family, and she had a healthy sibling (**Fig. 1Ai**). The patient’s mother had elevated serum cholesterol levels (total cholesterol: 291 mg/dl; LDL: 194 mg/dl; HDL: 82 mg/dl) and normal triglyceride levels (77 mg/dl). On the maternal side of the family, there was also a relative who had elevated serum cholesterol levels and passed away from a myocardial infarction before the age of 50.

The pregnancy was complicated by placental abruption at 32+2 weeks gestational age. She was born by an emergency C-section and spent 25 days in NICU due to prematurity. During the first years of life, she achieved all developmental milestones and was particularly athletic. When she was pre-teen, she sought medical attention due to difficulties with coordination and balance during dance lessons. She was diagnosed with complex HSP associated with ataxia that gradually deteriorated and limited her daily activities. She also developed slowly progressive dysarthria.

At her latest assessment, in her late teens (**Video 1**), she could walk independently for very short distances but required support for longer distances. Her gait was spastic and mildly ataxic. Her hearing and vision were normal. She was being treated with tizanidine and dantrolene and underwent regular lower limb botulinum toxin injections under sonographic guidance.

Physical examination revealed ankle clonus, increased tendon reflexes, and positive Babinski sign bilaterally. She had 3-4/5 paresis with pyramidal distribution in the lower limbs and 5-/5 paresis in the small muscles of the hand. She had mild gaze-evoked nystagmus, saccadic pursuit, mild dysarthria, mild dysmetria, and mild dysdiadochokinesis. Sensory testing was normal, and cognition was unimpaired. Her Scale for the Assessment and Rating of Ataxia (SARA) score was 14, and her Spastic Paraplegia Rating Scale (SPRS) score was 22. She had a high arched palate and a few white hairs on her head. Her skin was normal with no signs of ichthyosis. Her growth parameters were normal.

Her brain MRI was normal between the ages of 11 and 15 years (**Fig. 1Bi** and **iv**). A repeat brain MRI a few years later showed mild cerebellar atrophy and slightly enlarged ventricles (**Fig. 1Bii** and **v**). The most recent scan, performed in her late teens, was not substantially different (**Fig. 1Biii** and **vi**). Cervical and thoracic spinal cord MRI was normal (Data not shown). Her metabolic investigations, mtDNA sequencing, respiratory chain analysis from muscle biopsy and an extensive neurotransmitters panel in cerebrospinal fluid were normal.

#### Family 2

The second proband’s family was from the same geographic area as Family 1. There were two affected siblings: a female aged between 46 and 50 years (the proband) and her brother aged between 41 and 45 years (**Fig. 1Aii**). There was no known consanguinity in the family. Of note, the mother was affected by ALS.

#### Proband

The proband (II:1 Family 2) was in her late forties at the last examination (**Video 2**). She was born by emergency C-section after an 8-month gestation, with a birthweight of 1,650 grams, and spent 15 days in an incubator due to prematurity. She had strabismus at birth but no other neonatal problems and was otherwise a healthy infant. She had a normal neurodevelopmental development. As a young child, she had no difficulty with sports, and during her first decade of life, she underwent bilateral surgeries for strabismus.

Beginning in her late teens, she gradually developed a slow and unsteady gait, and by adolescence, she required intermittent assistance to walk. On neurological assessment between the ages of 11 and 15 years, she had a spastic gait requiring intermittent support. She was mildly dysarthric and had right exotropia, limitations of upward gaze, saccadic pursuit, mild dysmetria and dysdiadochokinesia. There was 4/5 paresis of pyramidal distribution in the lower limbs, increased tone in the legs, increased tendon reflexes in the lower limbs, ankle clonus, and bilateral positive Babinski sign. Sensory testing and hearing were normal.

By the end of her first decade, she was using a wheelchair but could still walk slowly for short distances with bilateral support. The gait was severely spastic, and the dysarthria and appendicular ataxia were significantly worse.

By the end of her thirties, she was wheelchair-bound and no longer ambulant. She had spastic tetraparesis (1-2/5 in the lower limbs and 4/5 in the upper limbs), severe dysarthria, severe appendicular ataxia, severe limitation of upward gaze with some restriction of horizontal gaze, and mild rigidity. Mild talipes equinovarus was noted. Sensory function could not be formally assessed.

At the last follow-up, aged between 46 and 50 years, she was tetraplegic, anarthric, with upper and lower limb contractures. Despite the anarthria, there was evidence of substantial comprehension. Her SARA score was 40, and her SPRS score was 50.

In her first decade, her brain MRI reportedly showed mild atrophy of the cerebellar vermis, but no other significant findings. Her electromyogram/nerve conduction studies (EMG/NCS), EEG, brainstem auditory evoked potentials (BAEP), and visual evoked potentials (VEPs) were reported normal. Somatosensory evoked potentials (SSEP) showed a conduction abnormality between the cervical medullary junction and the cortex. There was pigmentary change in the retinal periphery with reduced electroretinogram (ERG) responses. Her metabolic investigations in blood and cerebrospinal fluid were normal.

#### Affected sibling

The affected brother was in his early forties at the last follow-up. He was born by emergency C-section after an 8-month gestation, with a birthweight of 1,800 grams. He developed strabismus at an early age but had no other neonatal problems and was otherwise a healthy infant. His neurodevelopment was normal and had no difficulty with sports as a young child. There was no noteworthy symptomatology before the age of 10 years, when he gradually developed a slow and unsteady gait. On neurological assessment pre-teen, he had a mild right esotropia, very mild talipes equinus, slightly brisk lower limb tendon reflexes and equivocal plantars. In his early teens, he had a mildly spastic gait but could walk independently. There was a 4+/5 paresis of pyramidal distribution in the lower limbs. He had increased tendon reflexes, more pronounced in the lower than upper limbs, ankle clonus, and bilateral positive Babinski sign. There was mild appendicular ataxia and mild talipes equinus. Sensory testing and hearing were normal.

In his thirties, he was wheelchair-bound and no longer ambulant. He had spastic tetraparesis (2/5 in the lower limbs and 4+/5 in the upper limbs), severe dysarthria, severe appendicular ataxia, limitation of upward gaze, saccadic pursuit, slow saccades and mild rigidity. There were talipes equinovarus. Sensory function was normal. Cognition appeared unimpaired.

At the last follow-up, in his early forties (**Video 2**), he had further deteriorated, with severe tetraparesis (1/5 in the lower limbs and 2-4/5 in the upper limbs) and more severe dysarthria (could only whisper words). Sensation and cognition still appeared largely unimpaired. He could no longer feed himself. His SARA score was 38, and his SPRS score 46.

His brain MRI in his thirties reportedly showed atrophy of the cerebellar vermis. In his early childhood, EMG/NCS, EEG, BAEP and VEP were reported normal. SSEP showed a partial interruption of conduction in the spinal cord, brainstem and/or brain itself. There was mild reduction in ERG amplitudes with no evidence of retinal pigmentation.

#### Mother with ALS

Before her sixties, the mother was examined neurologically, and no significant abnormalities were found. She first noticed a left foot drop between the ages of 61 and 65 years. Within a few months, the paresis progressed proximally in the ipsilateral lower limb and distally in the contralateral lower limb. In a few years, she had left upper and lower limb paresis, milder right upper limb paresis, bilaterally increased tendon reflexes, left Babinski sign, and atrophy of the small muscles of the left hand, but no bulbar signs. She had evidence of active denervation in two levels and fasciculations at a third level. A diagnosis of probable ALS was soon updated to definite ALS. Over the next 3 years, her clinical picture deteriorated as expected for typical ALS. She died before the age of 70, tetraplegic, anarthric, with gastrostomy feeding and ventilator support.

### Genetic analyses identify a novel mutation G100R for FITM2

Exome sequencing of probands identified unreported variants in the *FITM2* gene present in the individuals with *FITM2* compound heterozygosity. The variant NM_001080472:c.298G>C (p.Gly100Arg, or G100R) was detected in both families.

In Family 1, the additional variant was NM_001080472:c.545_551dup p.(His185LeufsTer70, or H185-fs). In Family 2, it was NM_001080472: c.538C>T p.(Arg180Ter, or R180-Ter). Mapping of genetic variation throughout the genome did not reveal large regions of homozygosity on chromosome 20 in the region of *FITM2*, suggesting the G100R variant likely arose independently in both families (**Supplementary Fig. 1**).

In Family 1, targeted Sanger sequencing of parental samples confirmed that each parent carried a heterozygous variant of *FITM2*, confirming that the variants were biallelic (*in trans*). Molecular analysis of the healthy sibling showed that they did not carry the parental variants.

The first *FITM2* allele, H185-fs, was a previously unknown frameshift variant absent in control databases (gnomAd exomes r2.1.1: mean coverage 79.6, median coverage 100). This variant involves a seven-nucleotide duplication from DNA position 545 to position 551, resulting in a frameshift mutation starting at His185 and leading to altered amino acids after the frameshift and a premature termination codon 70 amino acids downstream (**Fig. 2A** and **Supplementary Fig. 2**). Classified as a likely pathogenic variant according to ACMG guidelines, it meets the criteria, including a null variant in a gene where loss of function is a known mechanism of disease (PVS1) and its absence from controls (PM2). It is predicted to result in the deletion of a large portion of FIT2, including one of the two histidine residues that are crucial for its enzymatic activity^11^.

**Figure 2.**
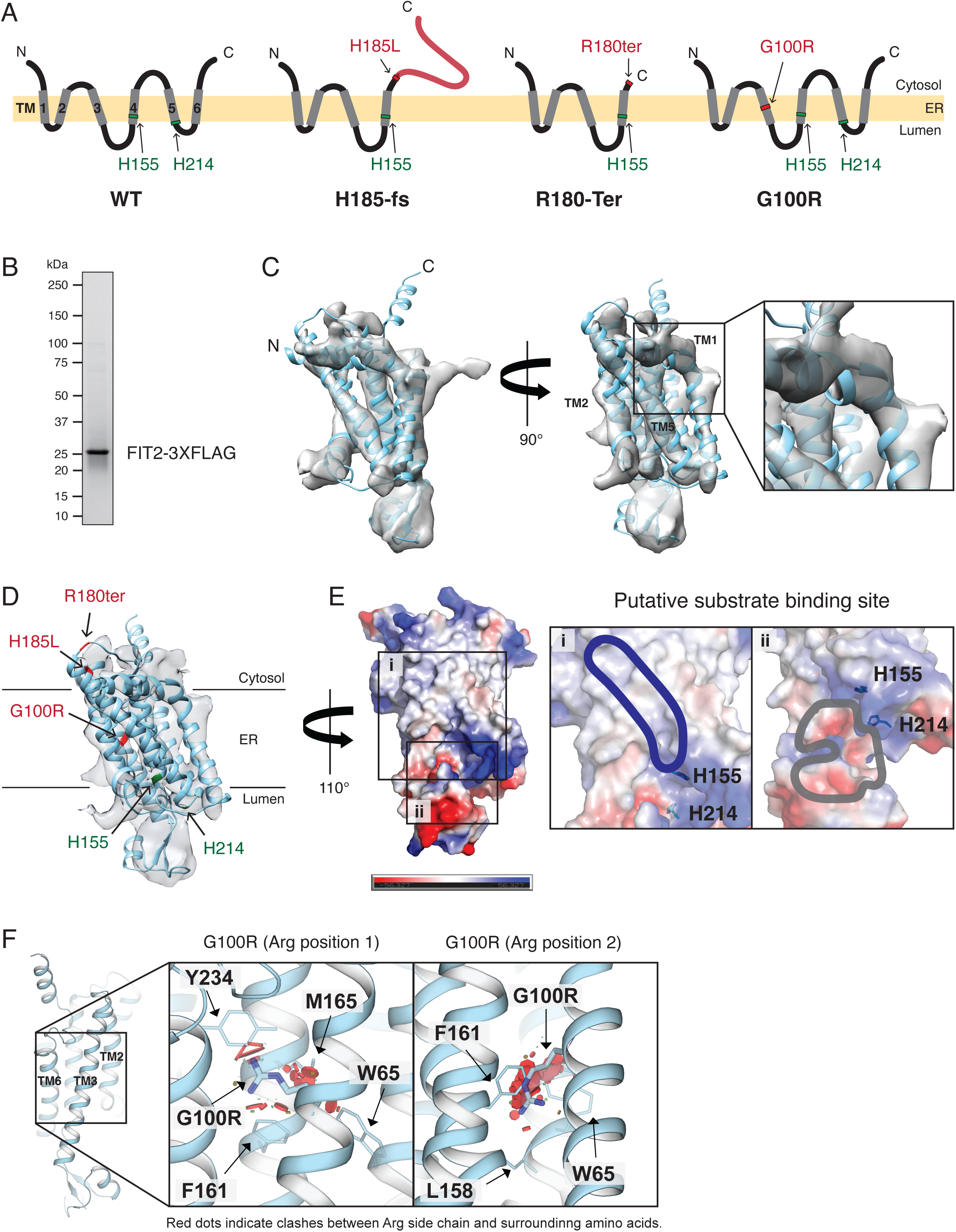
Genetic analysis and protein prediction. **(A)** Schematic Representation of the variants of the new families and the variants in the literature. FIT2 has six transmembranes (TMs). **(B)** SDS-PAGE of purified FIT2-3XFLAG. **(C)** FIT2 structure prediction (cyan) using AlphaFold with a cryo-EM density map (grey).**(D)** Three variants are highlighted in red and two catalytic histidines are highlighted in green. **(E)** Electrostatic potential surface of FIT2, with positive and negative electrostatic potentials are colored blue and red, respectively. Putative substrate binding site near the catalytic histidines is indicated by outlines. Putative binding sites of (i) acyl-chain and (ii) CoA are colored by blue and grey, respectively. **(F)** Visualization of G100R mutagenesis. Rotate 180 degree y-axis from (D). Representative clashes between G100R residue and other surrounding residues when the side chain of Arg towards up (Arg position 1, left) and down (Arg position 2, right). Red dots indicate possible clashes.

The second *FITM2* allele, p.Gly100Arg (G100R), is an extremely rare missense variant with a minor allele frequency of 0.000008% (gnomAD exomes r2.1.1: mean coverage 80.4, median coverage 100) with no homozygous carriers reported in the ExAC database. This variant results in the substitution of glycine with arginine at position 100 of the protein, predicted to be within the third transmembrane (TM) segment of FIT2 (**Fig. 2A** and **Supplementary Fig. 2**). It is classified as a variant of uncertain significance, according to ACMG guidelines, due to multiple lines of computational evidence, suggesting a deleterious effect on the gene or gene product (MetaRNN of 0.934, PP3), and its absence from controls (PM2).

Members of Family 2 also caried mutations in *FITM2*. Targeted Sanger sequencing of the ALS-affected mother revealed her as a sole carrier of the *FITM2* c.538C>T (p.Arg180Ter) variant; the father was unavailable for testing. We also screened the mother for repeat expansions in *C9ORF72* and other known genetic causes of ALS; however, no evidence was found to support these as the underlying cause of the disease.

The first allele of Family 2, *FITM2* p.Arg180Ter (abbreviated herein as R180-Ter), is a novel nonsense variant absent from control databases (gnomAd exomes r2.1.1: mean coverage 79.4, median coverage 100). It involves a change at position 538 where cytosine is substituted with thymine, generating a stop codon (TGA sequence) and a truncated protein at position 180 (**Fig. 2A** and **Supplementary Fig. 2**). Classified as likely pathogenic by the ACMG guidelines, it meets the same criteria as the p.His185LeufsTer70 variant. The second *FITM2* allele in the proband is G100R, as observed in Family 1.

### Mapping of the mutations by in silico and cryo-EM modelling of the FIT2 protein

To assess the impact of the G100R mutation, we sought to determine the structure of the FIT2 protein with cryo-electron microscopy (cryo-EM). We first expressed and purified the FIT2 protein to apparent homogeneity in buffer containing the detergent GDN (**Fig. 2B**). We analyzed the protein by single-particle cryo-EM and obtained EM densities for the protein (**Supplementary Fig. 3A**). The FIT2 particles were well-distributed and showed various orientations (**Supplementary Fig. 3B**), but the small molecular size (∼29.9 kDa) and its potential flexibility limited the resulting cryo-EM density map to relatively low resolution (∼6Å). To enhance our structural understanding, we combined the FIT2 cryo-EM map with AlphaFold predictions^28^. The model generated by AlphaFold, which posits six TM regions and one lumenal region, fit well into the experimental electron density map (**Fig. 2C** and **Supplementary Fig. 3C**), providing confidence in its accuracy.

The overall protein fold of FIT2 is similar to that of other proteins in the lipid phosphate phosphatases (LPP) protein family^29^. We detected structural similarities with several proteins: phosphatidylglycerophosphatase B (PgpB) from *Escherichia coli* (Z= 9.6)^30,31^, PgpB from *Bacillus subtilis* (Z= 8.5)^32^, and phosphatidic acid phosphatase type 2 (LpxE) from *Aquifex aeolicus* (Z=7.6)^33^. Unexpectedly, sphingomyelin synthase-related protein 1 (SMSr) from *Homo sapiens* (Z=7.8)^34^ also shares structural homology for the central helices (**Supplementary Fig. 4A**). The most similar protein, PgpB, is a member of the membrane-integrated type II phosphatidic acid phosphatase (PAP2) family of enzymes that are Mg^2+^-independent, have six TM domains, and dephosphorylate a broad variety of lipid and non-lipid substrates^35,36^. Members of the PAP2 family of enzymes share a signature sequence “KX_6_RPX_12–54_PSGHX_31–54_SRX_5_HX_3_D” that is divided into three motifs: C1, “KX_6_RPF”; C2, “PSGH”; and C3, “SRX_5_HX_3_D”^37^. The C1 domain is not found in FIT2^11^.

FIT2 contains six TM helices, with one of them containing the evolutionarily conserved catalytic histidine residues (**Supplementary Fig. 4B**) and a small lumenal domain of 35 amino acids (amino acids 119–153). Both the N-and C-termini are positioned on the cytosolic side of the membrane, similar to other LPP family members. The putative active site is located at the membrane-lumenal interface (**Fig. 2D**). A distinct groove near the catalytic site, likely facilitating substrate acyl CoA access, is also observed (**Fig. 2E**). The highly positively charged region is predicted to facilitate binding of phosphate moieties of coenzyme A, whereas the acyl chain is expected to reside within the elongated cavity of the protein within the ER membrane (**Fig. 2Ei**). Additionally, the lumenal domain may facilitate CoA binding (**Fig. 2Eii**).

Besides the central helices, other regions of FIT2 diverge from the structures of other LPPs. TM segments 2, 4, and 5 form the core of the TM region (**Supplementary Fig. 4C**), with TMs 1, 3, and 6 surrounding it; for PgpB, TMs 4–6 form the core, with TMs 1–3 surrounding it^30^. Notably, the TM1 of FIT2 is a helix bent ∼85 degrees (**Fig. 2C** and **Supplementary Fig. 4D**) and lacks homologous sequences. Bent helices can also be found in other LPP family proteins, but the angle and helix number are diverse. Additionally, PgpB contains a periplasmic domain inserted between TMs 3 and 4, which consists of four α-helices and may be involved in mediating substrate specificity^30^. In comparison, the lumenal domain of FIT2, also located between TMs 3 and 4, is shorter and consists of one α-helix and two β-strands that form a charged region (**Supplementary Fig. 4E**). This lumenal domain lacks the conserved C1 KX_6_RPF sequence found in most LPP enzymes that is involved in substrate recognition^11^.

The FIT2 cryo-EM-based model enabled us to map the identified mutations onto the predicted FIT2 structure (**Fig. 2D**). The H185-fs mutant maps to the TM4 segment. In this variant, nucleotides 545–551 are duplicated causing a frameshift that introduces a stop codon 70 residues later (AA 254) (**Supplementary Fig. 2**). The His185 is mutated to leucine. In the R180-Ter variant, the c.538C>T mutation introduces a premature stop codon after TM4. Both the H185-fs and R180-Ter variants are predicted to result in loss of the crucial second catalytic histidine (**Fig. 2A** and **Supplementary Fig. 2**) and therefore an inactive enzyme.

The G100R mutation introduces an elongated and charged arginine side chain into TM3 (**Fig. 2D**). Structural analyses indicate that the arginine residue causes significant steric hinderances with neighboring residues on adjacent TMs (**Fig. 2F** and **Supplementary Fig. 3D**). None of 22 backbone-dependent and 81 backbone-independent predicted arginine side chain rotamers is in a stable position. Representative illustrations of this are shown in **Fig. 2F**. In one orientation, the arginine side chain clashes with several surrounding amino acids, including Trp65, Phe161, Met165, and Try234. In an alternative orientation, it clashes with Trp65, Leu158, and Phe161. In most conformations, the position analogous to the G100 is occupied by either a glycine or an alanine (**Supplementary Fig. 4F**).

### FIT2 G100R mutant results in reduced FIT2 protein levels

We obtained human dermal fibroblasts (HDFs) from controls and patients (III:3, III:4, III:6 from family 1 and III:2 and III:3 from family 2) and measured the *FITM2* mRNA and encoded protein levels. *FITM2* mRNA levels were similar for patients and controls (**Fig. 3A**), indicating that alterations in mRNA levels are not the primary cause of the phenotypic differences in patients. However, FIT2 protein levels in patients were 75– 80% less than in control samples (**Fig. 3B**). The detected protein was of full length, indicating that the reduced amount of protein was likely from the G100R allele and not from one of the frameshift truncations. We hypothesized that the G100R substitution destabilizes the protein and causes its degradation. However, blocking proteasomal degradation with MG-132 did not significantly increase FIT2 protein levels (**Supplementary Fig. 5**). This suggests FIT2 G100R either is degraded through a mechanism not blocked by this proteasome inhibitor or that the mutation reduces efficiency of FIT2 translation.

**Figure 3.**
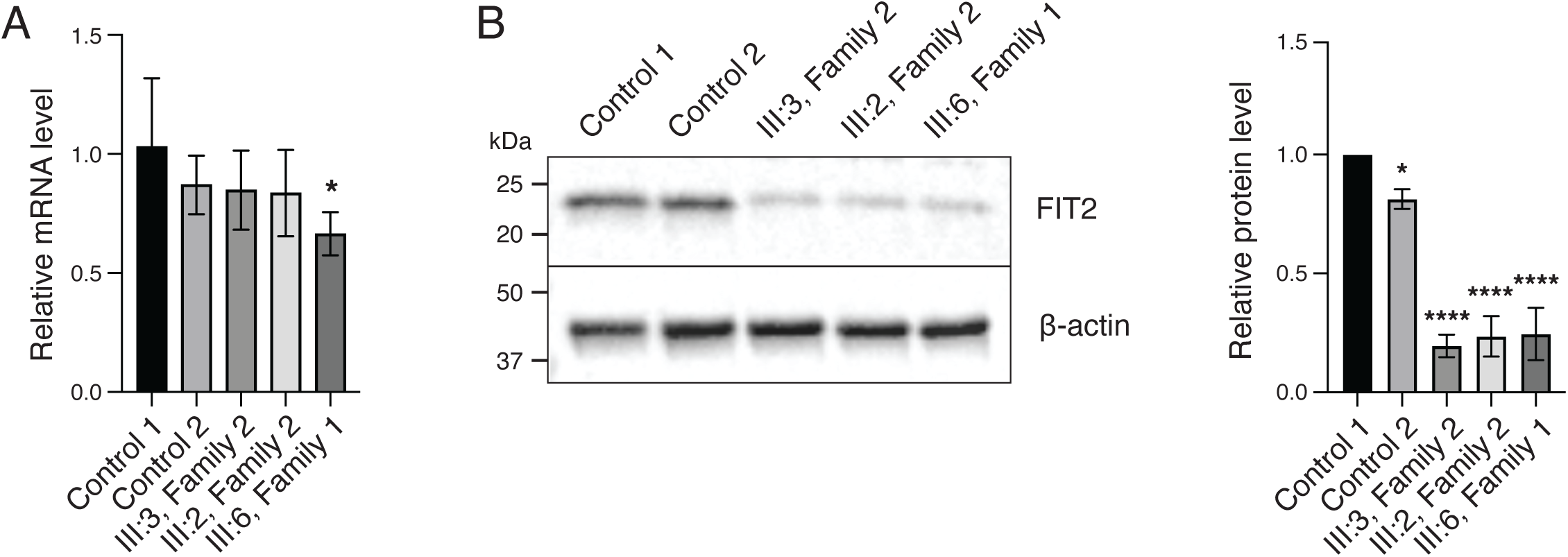
Reduced levels of the FIT2 protein in affected patient fibroblasts. **(A)** qRT-PCR analysis of *FIT2* transcript levels indicates that there are no significant changes between controls and patients. **(B)** Immunoblot analysis and quantification of FIT2 in human fibroblasts. Relative protein levels were normalized to β-actin. Data represent mean ± SD, n = 3/cell line, *p <0.05, **p < 0.01, ***p < 0.001, and ****p < 0.0001, vs. control 1, one-way ANOVA followed by post-hoc Dunnett’s multiple comparisons.

### FIT2 G100R possesses acyl-CoA diphosphatase activity

Although the G100R mutation causes reduced levels of the FIT2 enzyme, the mutant enzyme may still be active. We therefore determined the effect of the G100R mutation on FIT2’s specific activity as a fatty acyl CoA diphosphatase^11^. We overexpressed WT, H185-fs, G100R, and catalytically inactive mutant (HHAA) of FIT2 with C-terminal FLAG tag in Expi293F GnTI^−^ cells optimized for protein production, and isolated microsomes for enzymatic assays. We found that the expressed G100R mutant protein was present at lower protein amounts (∼20%) than WT or the HHAA mutant proteins (**Fig. 4A**). Assays of microsomes for acyl-CoA diphosphatase activity (measured by oleoyl phosphopantetheine production) revealed that FIT2 G100R exhibited less activity than the WT protein (**Fig. 4B**), at levels commensurate with its protein expression. These results indicate that G100R does not greatly reduce FIT2 activity, but instead reduces the amount of expressed protein.

**Figure 4.**
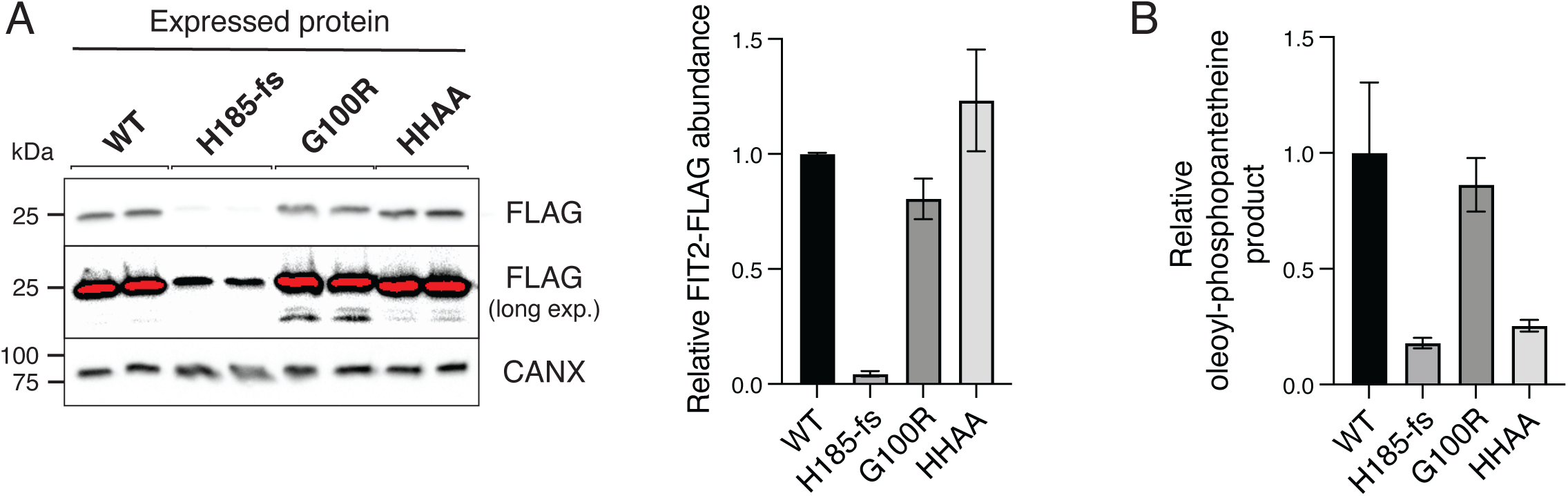
Biochemical analysis of patient FIT2 mutations indicates that G100R does not reduce FIT2 specific activity. FIT2-FLAG constructs were overexpressed in Expi 293F GnTI-cells. **(A)** Immunoblot analysis and quantification of FLAG-FIT2 in microsomal protein. **(B)** Acyl-CoA diphosphatase activity in FIT2-overexpress-ing microsomes used in A. Each reaction contained equal protein amounts and 25 μM oleoyl-CoA containing 0.1 μ Ci [^14^C]-oleoyl-CoA. Activity was measured as production of oleoyl-phosphopantetheine after 10 minutes and was normalized to PE (n=2/cell line). The H185-fs and HHAA have some background signals.

### FIT2 G100R is a hypomorphic allele in cellular assays

To further assess the function of the FIT2 G100R enzyme in cells, we expressed FIT2 WT and mutant enzymes from the AAVS1 “safe harbor” locus in FIT2 knockout (FIT2KO) SUM159 mammary carcinoma cells^11^. A construct encoding the H185-fs variant yielded no stably transfected cells, likely owing to the inherent instability of the H185-fs protein and was not included in subsequent experiments. For the G100R mutation, two clonal populations of WT and GFP-FIT2 G100R with variable FIT2 enzyme expression were isolated for analyses. Complementation with GFP-FIT2 G100R generated lower levels of FIT2 protein than that with the WT protein (GFP-FIT2 WT) (**Fig. 5A**). Consistently, fluorescence-based assays also showed reduced protein expression of the mutant protein (**Fig. 5B**, and data not shown), confirming the findings of reduced protein expression levels in patient fibroblasts.

**Figure 5.**
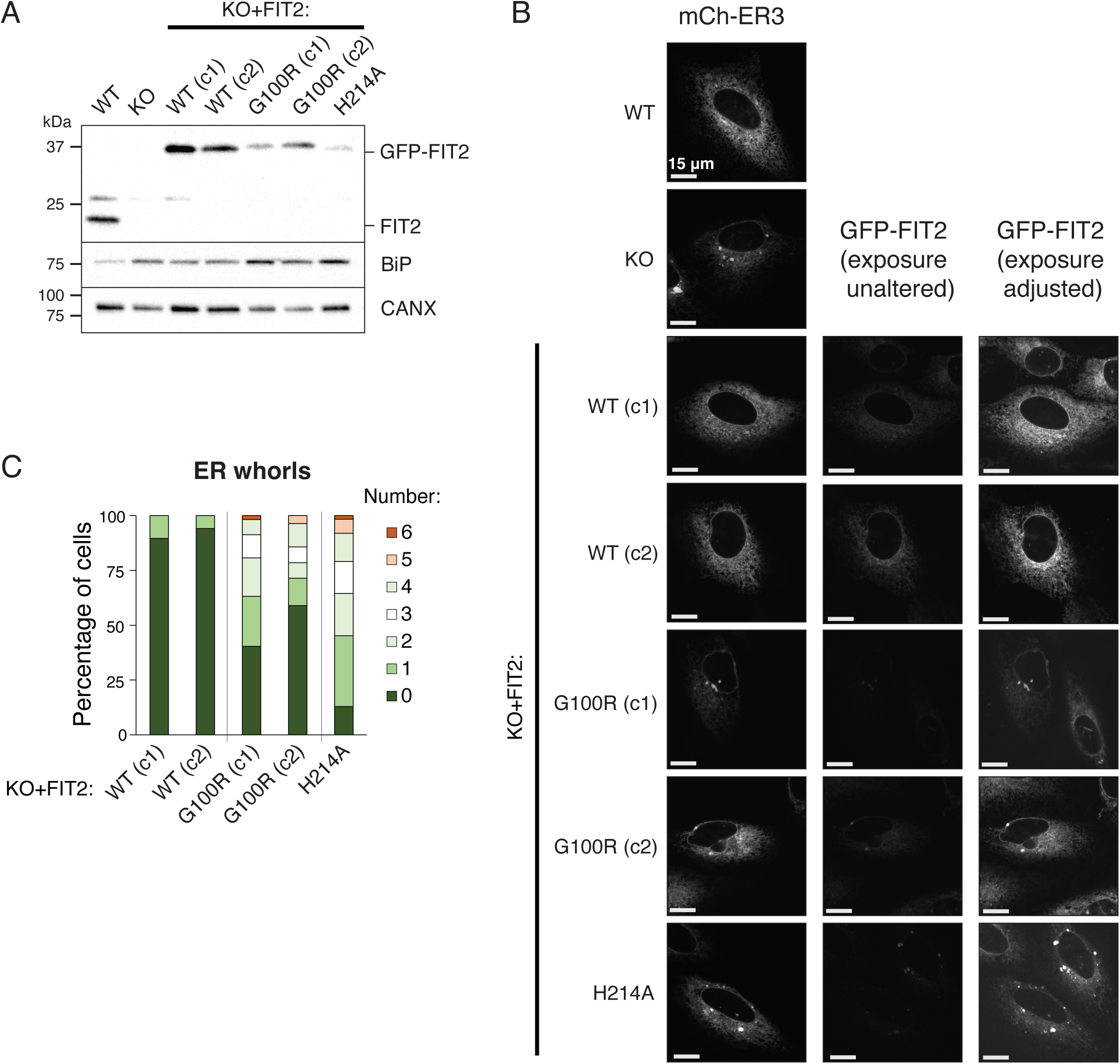
Molecular analysis in cultured mammalian cells demonstrates that G100R-FIT2 is a hypomorph. **(A)** Immunoblot analysis and quantification of FIT2 and BIP protein levels in whole-cell lysates. WT and G100R was reintroduced to FIT2KO SUM159 cells using the AAVS1 safe harbor locus. Single clones were isolated, and two clones for each genotype were used for subsequent analyses. **(B)** Confocal imaging of ER marker (mCherry-ER3) and GFP-FIT2 in WT, FIT2KO, KO cells with GFP-FIT-WT, -G100R and -H214A reintroduced. **(C)** Quantification of ER whorl number based on GFP-FIT2 signal in 40-60 cells per group.

FIT2KO cells exhibit alterations in ER morphology, including ER whorls^6^. We found that add-back of GFP-FIT2 WT rescued the ER morphology effect as expected, as visualized by the ER marker mCherry-ER3 and GFP-FIT2. In contrast, and consistent our previous report^11^, add back of catalytically dead GFP-FIT2 H214A resulted in cells with numerous ER whorls. Add-back of GFP-FIT2 G100R partially rescued the ER phenotype, with cells exhibiting ER whorls that were intermediate in terms of size and number (**Fig. 5B** and **C**).

FIT2 knockout cells exhibit signs of mild ER stress, apparent for instance in the increased expression of two unfolded protein response (UPR) target genes^12^. To test if rescue of ER stress accompanied the partial rescue in ER morphology, we measured transcript levels of UPR targets, BiP and Atf3 (**Supplementary Fig. 6**). GFP-FIT2 G100R only partially complemented the UPR induction phenotype, and this trend was maintained when measuring the levels of BIP (**Fig. 5A**). Overall, these experiments indicate that the FIT2 G100R variant results in less FIT2 enzyme than FIT2 WT, and that this reduction in FIT2 protein levels results in ER dysfunction in a human cell line.

The yeast *Saccharomyces cerevisiae* has orthologues for FITM2, such as *SCS3*, and deletion of *SCS3* results in numerous phenotypes, including inositol auxotrophy, that can be rescued by expression of the human protein^38^. We therefore tested whether the mutant proteins could rescue this phenotype in yeast. In agreement with our previous report, expression of WT human FIT2—but not catalytically inactive FIT2 (H155A)—rescued inositol auxotrophy in *scs3*Δ cells (**Supplementary Fig. 7A**). Further, we found that FIT2 H185-fs could not rescue this phenotype, whereas G100R partially rescued inositol auxotrophy.

Immunoblotting indicated that decreases in GFP-FIT2 protein levels correlated with reductions in FIT2 function (**Supplementary Fig. 7B**). We investigated GFP-FIT2 protein localization in yeast with fluorescence microscopy and determined that, while WT and FIT2 H155A localized to the ER and vacuole, H185-fs and G100R were restricted to the ER (**Supplementary Fig. 7C**). These imaging studies also allowed visualization of ER morphology. In agreement with previous studies^11^, we found that FIT2 WT expression yielded normal cortical and nuclear ER morphology, absent of “whorls”, but FIT2 H155A was unable to rescue the abnormal ER morphology (**Supplementary Fig. 7C and D**). FIT2 H185-fs also did not reduce ER whorl number or size, while G100R partially rescued both these ER morphology defects.

## DISCUSSION

In this study, we report a previously unknown hypomorphic allele of *FITM2* that is linked to HSP. This phenotype of *FITM2* mutations contrasts with those reported for subjects with homozygous (or compound heterozygous) null alleles that give rise to a deafness and dystonia syndrome, known as Siddiqi syndrome ^24–27^. The reported allele, G100R, was found in two distinct kindreds, and in each case combined with a null allele. Biochemical analysis of the effects of the G100R variant, as well as data from patient fibroblasts, cultured mammalian cells, and in yeast cells, all indicate that the protein retains activity but is expressed at lower levels than the wild-type protein, consistent with the allele being hypomorphic.

Notably, the hypomorphic allele apparently results in HSP rather than Siddiqi syndrome found with complete FIT2 deficiency. Thus, our studies add *FITM2* mutations to known genes that can cause a spectrum of neurodegenerative disorders. All the cases of *FITM2* deficiency in the literature (n=8) ^24–27^ describe early onset of symptoms (<1 year) (8/8), developmental delay (8/8) and failure to thrive (8/8), sensorineural deafness (8/8, described as the 1^st^ symptom in five affected children) associated to a progressive complex dystonia phenotype (6/8), ichthyosis-like features (6/8), sensory disturbances (5/8), and possible cognitive impairment (7/8) (**Table 1**). The reported cases displayed no spasticity, and only one of them showed an ataxic gait and was, interestingly, the only case associated with missense homozygous variants^25^.

In comparison with the classical described phenotype, the new cases we report present an early-onset progressive complex HSP associated with cerebellar ataxia. The probands from the two new families exhibited a later onset of the disease around the end of the first decade of life, with no prior neurodevelopmental delay and no failure to thrive. Deafness, dystonia, skin lesions, or neuropathy were not observed at the time of the most recent clinical follow-up assessment (in the late teens, early forties and late forties). MRI findings described in the reported cases were not uniform; however, both of our newly reported families showed cerebellar vermis atrophy and enlarged ventricles. Patient II:2 from family 2, in addition, presented with findings of Dandy-Walker spectrum, similar to the MRI findings described in the family reported by Shakir et al.^26^.

The previously reported cases of *FITM2* deficiency were subjects with homozygous (or compound heterozygous) null allelles^24–27^, with presumed null enzyme activity. These cases include families with homozygous E2X^24^ or homozygous M205X^27^. These mutations lack required catalytic histidine residues and are predicted to result in a complete loss of enzymatic function. In contrast, the G100R variant is a missense mutation that introduces a positively charged amino acid into a hydrophobic transmembrane segment but preserves both catalytic motifs of the enzyme. This allele resulted in lower amounts of enzyme that possess relatively normal enzymatic specific activity (i.e., activity levels correlate with the protein levels). The cause of the reduced amount of enzyme is unclear. Most likely, the amino acid substitution disrupts the enzyme folding and leads to accelerated degradation. However, we were unable to block enzyme degradation with the proteasome inhibitor MG-132. Nonetheless, inhibiting G100R degradation or stabilizing the mutant protein, analogous to strategies for the F508del mutation in cystic fibrosis, may be of benefit for patients suffering from HSP with this mutation.

By combining a low-resolution electron density map with AlphaFold predictions, our study provides structural insight into the FIT2 enzyme and its catalytic mechanism. As reported^11^, FIT2 is similar to other LPP enzymes and, in particular, to the PGP2 family of mainly membrane-integrated phosphatases^30,31^. Like other members of this enzyme family, FIT2 has two conserved catalytic histidine residues, which for FIT2 are predicted to reside at the ER-lumenal interface and are required for enzymatic activity^11,22^. However, FIT2 lacks the C1 KX_6_RPF sequence found in most LPP enzymes that is likely involved in substrate recognition. The FIT2 structure indicates that this region of the enzyme is part of a lumenal domain and suggests that it differs from typical family members. This difference in the substrate recognition region is consistent with biochemical data, indicating that FIT2 can utilize acyl-CoAs as substrates^11^. Supporting this further, the enzyme possesses a positively charged region near the catalytic site that may bind to phosphate moieties of coenzyme A. The FIT2 lumenal domain also exhibits a negatively charged region that is predicted to lie in the lumen of the ER. The function of this domain and whether it is involved in substrate recognition are unclear.

This study has limitations. First, it is a case series with a small sample size (three affected patients). Additionally, there is only a short follow-up time in Family 1, and additional phenotypic features may emerge over time. However, the long-term follow-up of the affected siblings in Family 2 (more than 30 years) is a strength, helping to differentiate the characteristics of this newly identified phenotype from the classical reported syndrome. The phenotype associated with homozygous G100R variants remains unknown and is likely very rare. Finally, future research should include larger sample sizes and longitudinal studies to validate and extend these findings.

## SUBJECTS AND METHODS

### Patient identification and clinical assessments

Both families lived in the same geographical area and are seemingly unrelated. The index case (II:2, Family 1) was referred to a specialized hospital at the at the end of her first decade due to suspicion of hereditary spastic paraplegia (HSP). After confirmation of the diagnosis, a genetic study was initiated to identify the causative gene of the disorder. Initial whole exome sequencing (WES) analysis failed to reveal any pathogenic/likely pathogenic variants or variants of unknown significance related to spastic paraplegia or ataxia genes. Cytogenetic analysis and array comparative genomic hybridization analysis by International Standard Cytogenomic Array Consortium oligonucleotide array 60K (Agilent) were normal. Additional testing included triplet expansion analysis for Friedreich ataxia and spinocerebellar ataxias type 1, 2, 3, 6, and 7, as well as mitochondrial DNA sequencing and deletions/duplications analysis in peripheral blood, all yielding normal results.

A revised WES analysis after 18 months, revealed two variants in compound heterozygosity in a newly recognized morbid gene *FITM2* (NM_001080472) encoding the fat storage-inducing transmembrane protein 2. Considering the gene’s function and its putative involvement in lipid droplet biogenesis^14–18^ and endoplasmic reticulum homeostasis^12,13^, further investigation was warranted to ascertain if additional cases of HSP harboring biallelic variants in *FITM2* existed.

Apart from the FITM2 gene variants, a heterozygous known pathogenic variant in the *LDLR* gene NM_000527:c.858C>A p.(Ser286Arg) was also identified as a secondary finding.

We adopted a genotype-first approach, meticulously analyzing extensive DNA sequence datasets from prominent diagnostic and research genetic laboratories. These datasets encompassed contributions from various entities, including Queen Square Genomics, the 100,000 Genomes Project, and ClinVar, supplemented by the utilization of GeneMatcher.

Subsequently, we identified two new cases presenting an HSP phenotype and harboring biallelic FITM2 variants, case III:2 and case III:3 from Family 2. Collaborating clinicians were contacted, and since both families resided in the same geographical area, they were able to undergo reassessment at the specialized hospital. Clinical features, complementary tests, and follow-up clinical data were collected from both families.

### Molecular genetic analyses

In both families, WES was performed on genomic DNA extracted from peripheral blood (QIAamp DNA Blood Kit) and captured with SureSelect V6+UTR-post technology (Agilent Technologies, Santa Clara, CA, USA). Sequencing was performed on an Illumina HiSeq 4000 sequencer (Illumina, San Diego, CA, USA) with 100 bp paired-end reads. Sequence reads were aligned to the GRCh37 (hg19) human genome reference. Read mapping, variant calling and variant annotation were performed using the software BWA-0.7.12, GATKv3.4.0 and SnpEff_v4.1g, respectively. Variant assessment involved evaluating frequency in population databases (including gnomAD), predicted pathogenicity *in silico*, and phenotypic overlap.

Candidate variants in both families were confirmed after filtering and interpretation according to the American College of Medical Genetics and Genomics and the Association for Molecular Pathology (ACMG-AMP) guidelines. Segregation analysis was performed by Sanger sequencing on an ABI 3500xL (Life Technologies, Carlsbad, CA).

Since both families were of the same origin and shared one of the two variants in heterozygosity, an analysis of regions of homozygosity from sequencing data was conducted to detect a potential founder effect in both families using the software available at https://automap.iob.ch/.

### In silico modelling of the FIT2 protein

The conservation of Gly100 was determined through alignment of the FIT2 protein sequence across different species using Clustal Omega (https://www.ebi.ac.uk/jdispatcher/msa/clustalo) and protein BLAST (https://blast.ncbi.nlm. nih.gov/Blast.cgi).

A predicted model of human FIT2 (UniProt ID: Q8N6M3) was generated by AlphaFold^28^. A hydrophobicity plot was used to determine a thermodynamically favorable orientation of FIT2 within the ER bilayer. The position of the new amino acid changes within the structure was estimated using UCSF ChimeraX^39^, Dynamut2 (https://biosig.lab.uq.edu.au/dynamut2/), and PyMOL (The PyMOL Molecular Graphics System, Version 2.5.4 Schrödinger, LLC.). For the figure preparations, UCSF Chimera X and PyMOL were used.

### Cryo-EM sample preparation and data acquisition

The FIT2 protein was expressed in the human embryonic kidney (HEK)293S GnTI^-^ suspension cells (ATCC), cultured in FreeStyle 293 Expression Medium (Thermo Fisher Scientific, 12338026) with 2% (v/v) FBS at 37°C under 8% CO_2_. When cell density reached ∼2.5M mL^-1^, the pCMV14-FIT2-3xFLAG plasmids were transfected into the cells. For 1 liter of cell culture, 1 mg of plasmids were pre-mixed with 3 mg of linear polyethyleneimine (Polysciences, 23966) in 40 mL of Opti-MEM medium for 30 mins at room temperature before transfection. After 18 hours of shaking incubation at 37°C, 10 mM sodium butyrate was added, and the growth temperature was lowered to 30°C to boost protein expression. After 48 hours of treating sodium butyrate, the cells were harvested by centrifugation at 550 g and resuspended in lysis buffer (20 mM Tris, pH 8.0, 150 mM NaCl, 1 mM PMSF, 10% (v/v) glycerol) supplemented with cOmplete protease Inhibitor Cocktail tablet, EDTA-Free. Membranes were solubilized by adding 2% (w/v) GDN (Anatrace, GDN101) at 4°C by gentle agitation for 1 hour followed by centrifugation at 14,000 rpm for 1 hour to remove insoluble materials. The supernatant was subsequently incubated with anti-FLAG M2 resin (Sigma-Aldrich, A2220) for 1 hour. The resin was then packed onto a gravity-flow column (Bio-Rad, 7321010) and washed with 10 column volumes (CV) of buffer A (20 mM Tris pH 8, 150 mM NaCl, 0.02% (w/v) GDN). The FIT2 protein was eluted by 5CV of elution buffer (20 mM Tris, pH 8, 150 mM NaCl, 0.02% (w/v) GDN, 0.150 mg mL^-1^ 3xFLAG peptide). The eluted protein was concentrated and further purified by size-exclusion chromatography (SEC) on a Superdex 200 Increase column (Cytiva Life Science, 28-9909-44) equilibrated with buffer A. Peak fractions were pooled and concentrated to 8–10 mg mL^-1^ for cryo-EM analysis. For cryo-EM analysis, the concentrated protein was incubated with 3 mM Fos-Choline-8 (Anatrace, F300F). All grids were prepared with FEI Vitrobot Mark IV (FEI Thermo Fisher) at 4°C and 100% humidity. 3.5 μL of the sample was applied to fresh glow discharged Quantifoil R1.2/1.3, Cu 400 mesh (Electron Microscopy Sciences, Q4100CR1.3).

Cryo-EM data were collected with a Titan Krios microscope (FEI) operating at 300 keV equipped with a K3 summit direct electron detector (Gatan). Movie datasets were collected with SerialEM in super-resolution mode at 29,000x with a super-resolution pixel size of 0.413Å. Each movie contained 60 frames over 3 s exposure time, using a dose rate of ∼15 e^-^Å^-2^s^-1^, resulting in a total dose of ∼66 e^-^Å^-2^, and the defocus range was -0.6 to -1.7 μm.

A total of 4,058 and 4,545 super-resolution movies were collected. All two datasets were processed similarly. The movies were normalized and motion corrected using the patch-motion-correction algorithm by cryoSPARC (v4.4.0)^40^. Blob-based autopicking in cryoSPARC was implemented to select initial particles. After several rounds of 2D classification, selected particles were used for training in Topaz particle-picking pipeline^41^. Particles picked by Topaz were subjected to several rounds of 2D classification for clean-up and selected final particles from each dataset (499,378 and 576,398 particles) were combined to perform the *ab initio* procedure in cryoSPARC. Three parallel *ab initio* reconstructions jobs were performed using identical settings, and particles from the class showing better features were selected from each job and combined for the next *ab initio* procedure^42^. After several rounds of *ab initio* procedures, the final 95,285 particles were used for the acceptable FIT2 map with six transmembrane features.

### Plasmid construction

Plasmids in this paper are described in **Supplementary Table 1**. pMB111-v1 and pMB111-v2 (expressing GFP-FIT2 H185-fs and GFP-FIT2 G100R, respectively) were derived from pMB111 (expressing GFP-FIT2 WT) via cloning of FIT2 mutant sequence using EcoRI-HF and XhoI sites. pMB65-v1 and pMB65-v2 (expressing GFP-FIT2 H185-fs and GFP-FIT2 G100R, respectively) were derived from mMB65 (expressing GFP-FIT2 WT) via cloning of FIT2 mutant sequence using AfeI and PmeI sites. pMB82-v1 and pMB-v2 (expressing FIT2 H185-fs-3xFLAG and FIT2 G100R-3xFLAG, respectively) were derived from mMB82 (expressing FIT2 WT-3xFLAG) via site-directed mutagenesis. For pMB82-v1, PCR primer pairs GACTTCCTCCCCAGAGCTGGAGAATCTGTACTTCCAG and CTGGAAGTACAGATTCTCCAGCTCTGGGGAGGAAGTC were used. For pMB82-v2, PCR primer pairs GCACCCTGCTTGTGCGCACGGCCATCTGGTAC and GTACCAGATGGCCGTGCGCACAAGCAGGGTGC were used. Mutagenized plasmids were checked by sequencing.

### Yeast growth conditions and growth assays

All yeast strains were derivatives of the BY4741 strain, as described^6^ (**Supplementary Table 2**). *Scs3*Δ were transformed by a lithium acetate/single-stranded carrier DNA/polyethylene glycol procedure^43^. Cells were grown in synthetic complete (SC) medium containing 2% (wt/vol) glucose without uracil or leucine for plasmid selection. For drop assays, WT or *scs3*Δ yeast transformed with pMB111 (GFP-FIT2 WT), pMB111-v1 (GFP-FIT2 H185-fs) pMB111-v2 (GFP-FIT2 G100R), and pMB112 (GFP-FIT2 H155A) were grown overnight in SC without uracil liquid culture to stationary phase and spotted in 10-fold serial dilutions onto synthetic medium plates without uracil with or without inositol (2 µg/µl). Images were obtained after 2 days of growth at 37°C.

### Cell culture

Human SUM159 breast carcinoma cells (gift from T. Kirchhausen) were maintained in DMEM/F-12 GlutaMAX medium (Life Technologies, 10565-042) supplemented with 5% FBS, 1 mg/mL hydrocortisone (Sigma-Aldrich, H0888), 5 mg/mL insulin (Cell Applications, 128-100), 10 mM Hepes, pH 7.0, 100 U/mL penicillin, and 100 mg/mL streptomycin. Transfection of plasmids was performed 24 h before experiments and was mediated with FuGENE-HD (Promega, E2312) according to the manufacturer’s instructions. Expi293F GnTI^-^ cells were grown in FreeStyle 293 Expression Medium (Thermo Fisher Scientific, 12338026) in suspension in according to the manufacturer’s instructions.

HDFs were cultured in DMEM (Thermo Fisher Scientific, 10-569-044) supplemented with 10% HI-FBS at 37°C under 5% CO_2_. HDFs were harvested when confluent. When needed, HDFs were treated with 10 µM MG-132 (Sigma-Aldrich, 474791) for 24 hours before harvesting.

### Generation of FITM2 patient mutation knock-in cells

To generate cell lines stably expressing GFP-FIT2 and GFP-FIT2 variants in FIT2-KO SUM159 cells, we used the AAVS1 safe harbor targeting method (System Biosciences). GFP-FIT2 WT, GFP-FIT2 G100R, GFP-FIT2 H185-fs, or GFP-FIT2 H214A donor constructs were cotransfected with a plasmid containing hCas9 plasmid and a gRNA-AAVS1-T2 plasmid using Fugene HD (Promega, E2312) according to the manufacturer’s instructions. Successfully transfected cells were selected for with puromycin treatment for 3 days, followed by single-cell FACS sorting on the basis of GFP signal. Transfection with FIT2 H185-fs plasmid pMB65-v1 did not produce any cells with GFP signal, likely because of instability of this protein variant as described in results and was not included in subsequent experiments. Positive clones were confirmed by fluorescence microscopy and western blotting to confirm protein expression of appropriate molecular mass. Cells were tested for mycoplasma contamination using PCR Mycoplasma Detection Kit (Applied Biological Materials, G238).

### RNA isolation and RT-qPCR

Total RNA was isolated from cultured SUM159 cells and HDFs using the RNeasy Mini Kit (Qiagen, 74104), according to the manufacturer’s instructions. RNA was reverse transcribed to obtain cDNA using the iScript cDNA synthesis kit (Bio-Rad Laboratories, 1708891). The qPCR was performed using Power SYBR Green Master Mix (Applied Biosystems for SUM159 cells and Thermo Fisher Scientific, 4368702 for HDFs) with a Bio-Rad CFX96 Real-Time System. The expression level was normalized to the housekeeping gene *CyclophilinA* and evaluated by comparative *C*_T_ value. Relative mRNA expression level was calculated using the 2^−ΔΔCT^ method. Sequences of the qPCR primers used are listed in **Supplementary Table 3**.

### Oleoyl-CoA flux measurement in microsomes

Cell transfection, microsomal preparation and flux measurements were conducted as described^6^. Expi293F GnTI^-^ suspension cells were transfected with plasmids pMB82 (FIT2 WT-3xFLAG), pMB82v1 (FIT2 H185-fs-3xFLAG) and, pMB82v2 (FIT2 G100R-3xFLAG) and, or pMB277 (FIT2 HHAA-3xFLAG) and 1 mg/ml polyethyleneimine (Polysciences, 23966). At 16 hours after transfection, 10 mM sodium butyrate (Sigma-Aldrich, 303410) was added to the cell culture, and at 48 hours after transfection, cells were harvested. The cell pellet was resuspended in lysis buffer (250 mM sucrose, 50 mM Tris-HCl, pH 7.4, 200 mM NaCl, 1 mM EDTA) supplemented with protease inhibitors (Roche). Cells were lysed using a Dounce homogenizer, and the cell lysates were centrifuged at 8,000 g for 10 min at 4°C to remove cell debris, followed by further centrifugation of the supernatant at 100,000 g for 60 min at 4°C to isolate microsomes. The pellet was resuspended in lysis buffer. For flux reactions, 100 µg of microsomes were mixed with reaction buffer (100 mM KAc, pH 7.0, 1 mM MgCl_2_, 20 mM NaCl, 0.625 mg/mL BSA) and adjusted to a final volume of 200 µL with lysis buffer. The assay was initiated through addition of 25 µM oleoyl-CoA containing [^14^C] oleoyl-CoA (American Radiolabeled Chemicals, 0527-50 µCi) as a tracer (0.1 µCi). Reactions were incubated at 37°C for 10 min and then stopped by addition of chloroform:methanol (2:1) and 2% phosphoric acid. The lower phase (containing lipid) was collected, dried under an air stream, resuspended in chloroform, and separated by TLC with a chloroform:methanol:water (65:25:4) solvent system to separate polar lipids. The TLC plates were exposed to a phosphor imager screen for 2 days and developed with the Typhoon FLA 7000 phosphor imager.

### Live-cell imaging and image processing

Microscopy was performed on a spinning disk confocal microscope (Yokogawa CSU-X1), paired with a Nikon Eclipse Ti inverted microscope with a 100 Apochromat total internal reflection fluorescence/1.4 NA objective (Nikon). Then 405-, 488-, or 561-nm laser lines were used for fluorophore excitation, and fluorescence was detected with a Zyla 4.2 Plus scientific complementary metal-oxide-semiconductor camera (Andor). Acquired images were processed and quantified manually with ImageJ software. For assessment of ER morphology, z-stacks of 3-micron depth with 300-nm spacing were acquired, and z-projections were obtained with four z-stacks. Whorl number and severity were manually quantified based on GFP-FIT2 signal from 20–30 cells per group (for yeast imaging) and 40–60 cells per group (for mammalian cell imaging).

### Antibodies and immunoblotting

Mammalian cell extracts were prepared for western blot analysis by lysing with lysis buffer (150 mM NaCl, 50 mM Tris-HCl, pH 7.4, 1% (v/v) Triton X-100 supplemented with a protease inhibitor cocktail), followed by centrifugation for 6 min at 16,000 g. The supernatant was denatured in Laemmli buffer at 37°C for 10 min. Protein extracts were separated on a 12% SDS-PAGE gel (Bio-Rad Laboratories) and transferred to a polyvinylidene difluoride membrane (Bio-Rad Laboratories). Yeast extracts were prepared for western blot analysis by lysing cells in 10% trichloroacetic acid, supplemented with protease inhibitor cocktail, bead vortexing at 4°C for 10 min, and centrifugation for 10 min at 16,000 g. The pellet containing precipitated protein was resuspended in Laemmli buffer containing β-mercaptoethanol and heated at 37°C for 10 min. Mammalian cell and yeast extracts were separated on a 4–15% SDS-PAGE gel (Bio-Rad Laboratories) and transferred to a polyvinylidene difluoride membrane (Bio-Rad Laboratories). Human FIT2 protein was detected using a custom rabbit polyclonal antibody (1:1000 dilution) with epitope corresponding to C-terminal 16 residues of FIT2. Generation of this custom FIT2 antibody was described.^6^ Other antibodies include anti-FLAG (Sigma-Aldrich, F1804, 1:1000), CANX (Cell Signaling C5C9, 1:500), Pgk1 (Abcam ab113687, 1:3000) and GFP (Roche 11814460001, 1:500).

Cultured HDFs were lysed in RIPA buffer (Thermo Fisher Scientific, 89900), supplemented with EDTA-free protease inhibitor cocktail (Roche, 04693132001) and 1% (v/v) Triton X-100. After incubating for 1 hour, cells were spun for 30 min at 20,000 g. The supernatant was then denatured in Laemmli sample buffer (Bio-Rad, 1610747). Proteins were separated on a 4–15% SDS-PAGE gel (Bio-Rad, 4568084) and transferred to a nitrocellulose membrane (Thermo Fisher Scientific, IB23001). Human FIT2 protein was detected as described, with β-actin (Santa Cruz, sc-47778) as a control. To evaluate the function of MG-132, we used an anti-ubiquitin antibody (Cell Signaling, 3936T).

### Statistics

Data in Figures 3, 4, 5 and Supplementary Figures 6 and 7 results were expressed as mean ± standard deviation (SD). Replicates are expressed as n= number of replicates/cell line. If not indicated differently, data were compared by 1-way ANOVA, followed by post-hoc Dunnett’s adjustment for multiple comparisons. A P value of less than 0.05 was considered significant, and asterisks in graphs denote the significance of P values comparing indicated group to control (*P < 0.05; **P < 0.01; ***P < 0.001, ****P < 0.0001). Bars with no asterisks indicate no significant difference from the designated control. GraphPad Prism (v8 and v10) was used to prepare graphs and perform statistical analysis.

## Data Availability

All data produced in the present study are available upon reasonable request to the authors

## Abbreviations

BAEP: brainstem auditory evoked potentials
EMG: electromyogram
ERG: electroretinogram
FIT2: fat-storage transmembrane protein 2
HSP: hereditary spastic paraplegia
NCS: nerve conduction studies
SARA: Scale for the Assessment and Rating of Ataxia
SSEP: somatosensory evoked potentials
SPRS: Spastic Paraplegia Rating Scale
UPR: unfolded protein response
VEP: visual evoked potentials
WES: whole exome sequencing

## Study approval

This study was conducted in accordance with the Declaration of Helsinki. Ethical approval for this study was obtained from the institutional review boards of University College London and the respective host institutions. The patients and their relatives provided written informed consent for the collection, storage, and publication of the clinical data, blood samples, and experimental results under the International Genomics Collaboration Study (IRAS 310045). All clinical photographs and videos appear with parental informed written consent.

## Data availability

The primary data sets that support the findings of this study are available from the corresponding author, upon request. Omics data will be posted on a public server.

## Author contributions

The fact that this article has four main authors reflects the best way to represent the collaborative work between different groups involved in the study discussed in this article. K.A. is the person who identified the gene reported as likely responsible for the phenotype and, together with A.S-V., has led the genetic and clinical characterization aspect. L.K. and L.M.B. are the ones who conducted the functional studies. M.K., G.Ko., R.V.F., T.C.W., H.H. and A.S-V conceptualized the study. G.Ko., M.K., G.Ka, A.D., and E.F. performed clinical assessment of the patients. A.S-V, M.K., G.Ko and H.H analyzed and interpreted the phenotype data. A.S. cultured and maintained skin biopsy-derived fibroblasts. D.M. was responsible for the bioinformatic processes of the included genetic data. A.S-V, S.E., K.A., and A.V. analyzed genetic data and performed Sanger sequencing. L.M.B. performed the enzymatic activity assays using microsomes, collected and analyzed confocal images, and performed yeast experiments. L.K. performed protein expression and purification, prepared cryo-EM grids, collected and processed cryo-EM datasets, analyzed protein structures, and measured mRNA and protein levels from HDFs. L.M.B., L.K., R.V.F., and T.C.W. analyzed and interpreted the data. A.S-V., L.M.B., L.K., R.V.F. and T.C.W. wrote the manuscript. All authors discussed the results and commented on the manuscript.

## Acknowledgments

We thank M. J. de la Cruz of the Structural Biology core Facility at the Memorial Sloan Kettering Cancer Center for help with data acquisition, T. Kirchhausen for providing the SUM159 cells, and G. Howard for editorial assistance. This work was supported by NIH grant R01GM141050 (to R.V.F).

L. M. B. was supported by the National Institute of Health Service Award T32 DK00747 and an American Heart Association Postdoctoral Fellowship A.S-V was supported by La Caixa Postgraduate Abroad fellowship.

---

**Figure S1.**
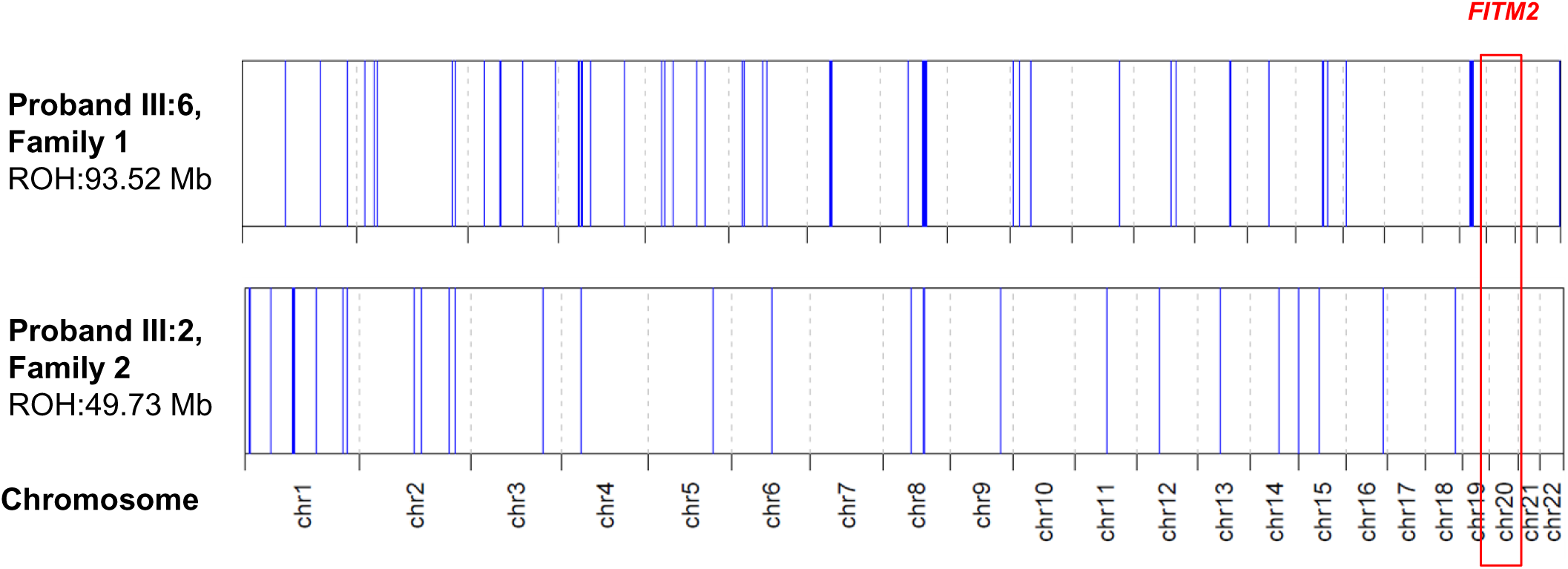
Homozygosity mapping. Autozygosity mapping using the AutoMap4 tool did not identify any common region of homozygosity (ROH) on chromosome 20, where the *FITM2* gene is located, when comparing the raw exome data (VCF files) from both probands. Family references are displayed on the left as a guide. Regions of homozygosity identified in single individuals are shown in blue, with their total genomic extension across autosomes (in Mb) reported below each family reference. Each chromosome location is delimited by dashed lines, and the location of chromosome 20 is marked with red boxes. GRCh38/hg38 = Genome Reference Consortium Human Build 38 Organism: *Homo sapiens* (human); Mb = megabase(s); ROH = region of homozygosity; Chr = chromosome.

**Figure S2.**
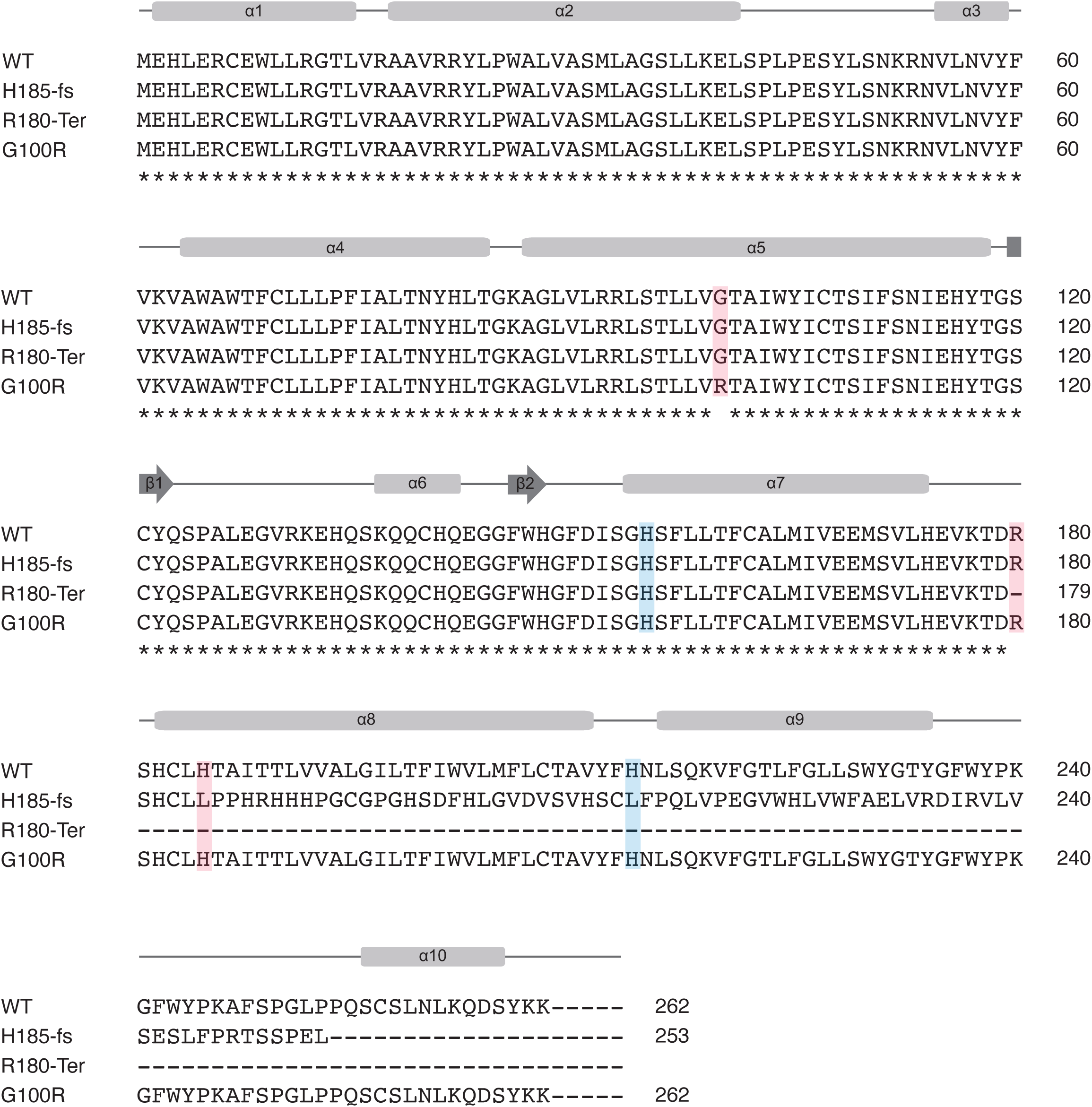
Protein sequence alignment of FIT2 WT and variants. The sequence alignment of FIT2 WT, H185-fs, R180-Ter, and G100R. Secondary structure elements based on FIT2 WT are shown as gray cylinders (helices) and arrows (beta-strand). Two catalytic histidines (H155 and H215) are highlighted in blue. Affected variants are highlighted in red. Molecular mass of each protein is 29.86 kDa (WT), 28.78 kDa (H185-fs), 20.43 kDa (R180-Ter), and 29.95 kDa (G100R).

**Figure S3.**
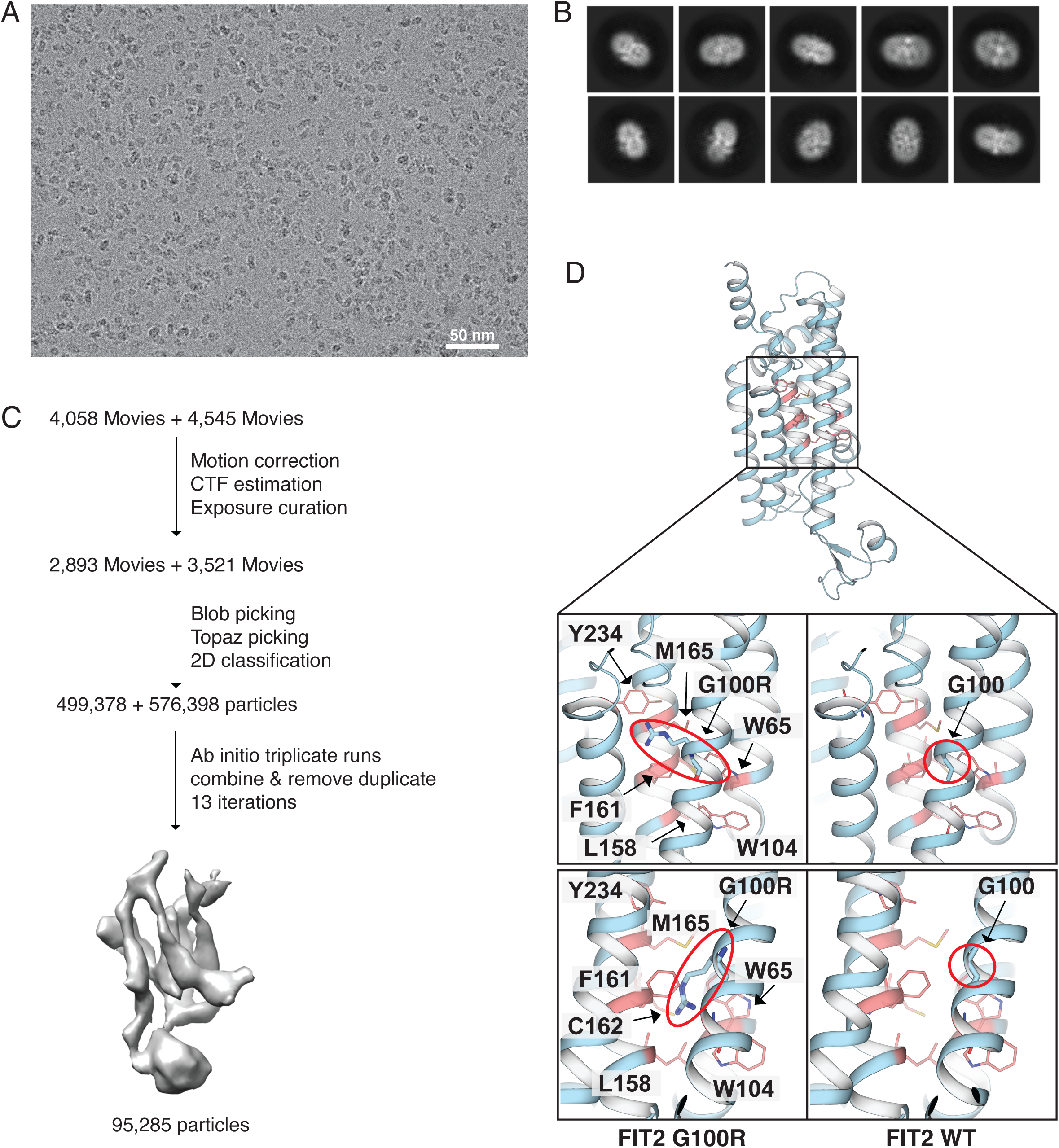
Cryo-EM analysis of FIT2. **(A)** The image shows a representative cryo-EM image of FIT2 particles. **(B)** Representative 2D class averages. **(C)** A workflow for cryo-EM data processing (see Methods for details). **(D)** The side chain of arginine clashes with many surrounding amino acids (e.g., Trp65, Trp104, Leu158, Phe161, Cys162, Met165, and Tyr234), which are colored pink, depending on the direction of the side chain. Red circles indicate side and main chains of G100R and G100.

**Figure S4.**
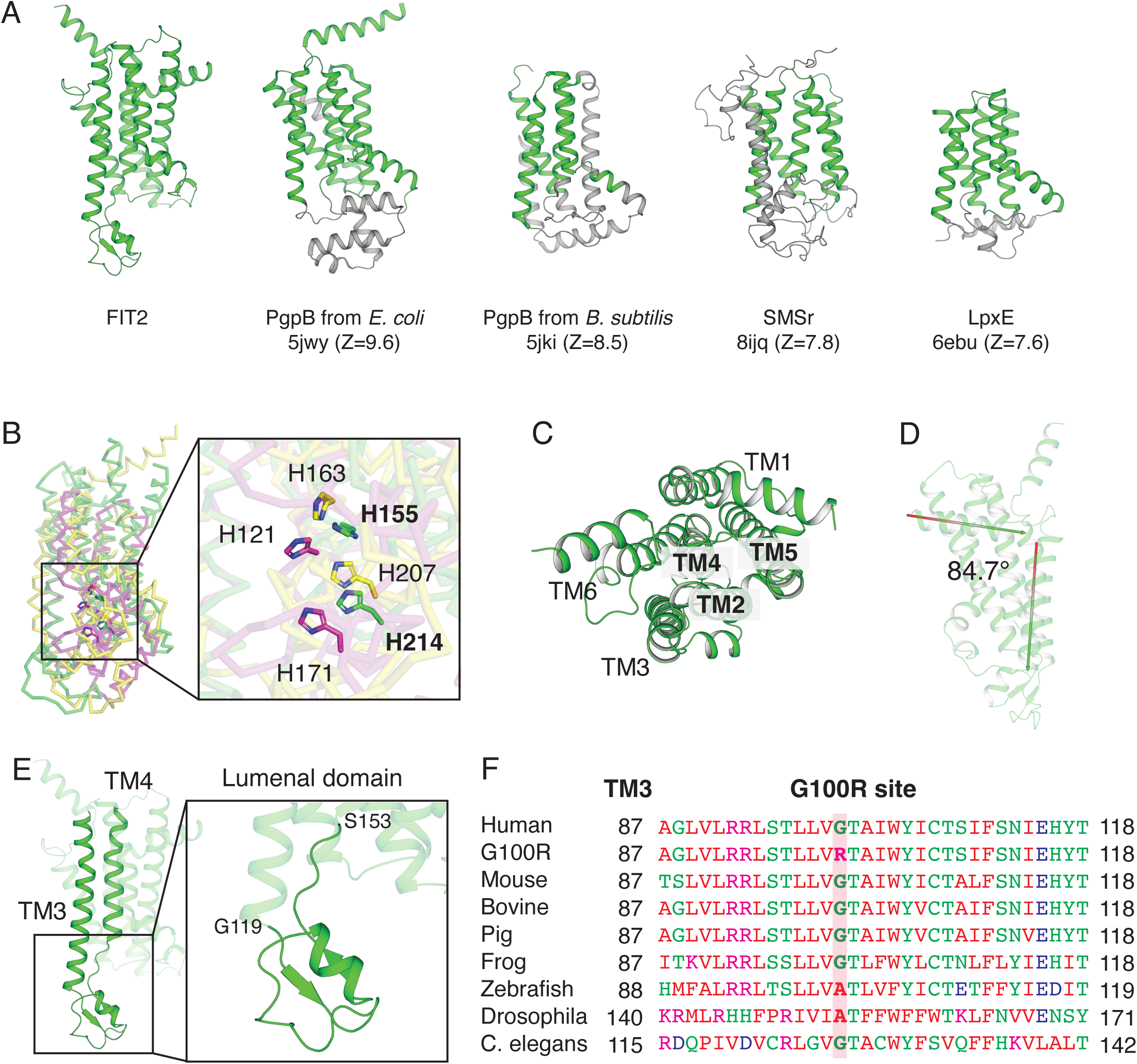
Structure comparison. **(A)** Ribbon diagrams comparing the overall structures of FIT2, PgpB, LpxE, and SMSr. PDB codes and Z-scores from the DALI server for each structure are shown below each structures. Structural regions that align with FIT2 are highlighted in green. **(B)** Structural overlay of FIT2 (green), PgpB from *E. coli* (yellow) and PgpB from *B. subtilis* (magenta) performed using the cealign tool in PyMOL. The catalytic histidines are similarly positioned and oriented in the same direction across the structures. **(C)** A periplasm view of the FIT2 structure, rotated 90 degrees around the X-axis from Fig. S4A FIT2 structure. The core TMs (TM2, 4, 5) are indicated in bold. **(D)** The degree of bent helix measured by AngleBetweenHelices method in PyMOL. **(E)** The lumenal domain (119-153) of FIT2. It contains one α-helix and two β-strands between TM3 and TM4. **(F)** Amino acid conservation across species at the position of the novel substitution (highlighted in red) in TM3. Red colors present small and hydrophobic, blue for acidic, magenta for basic, green for hydroxyl, sulfhydryl and amine residues. UniProt ID: human (Q8N6M3), mouse (P59266), bovine (A4IFN5), pig (B2LYG4), frog (Q6AX73), zebrafish (Q52KL1), *Drosophila* (Q9VRJ2), *C. elegans* (Q5CZ37).

**Figure S5.**
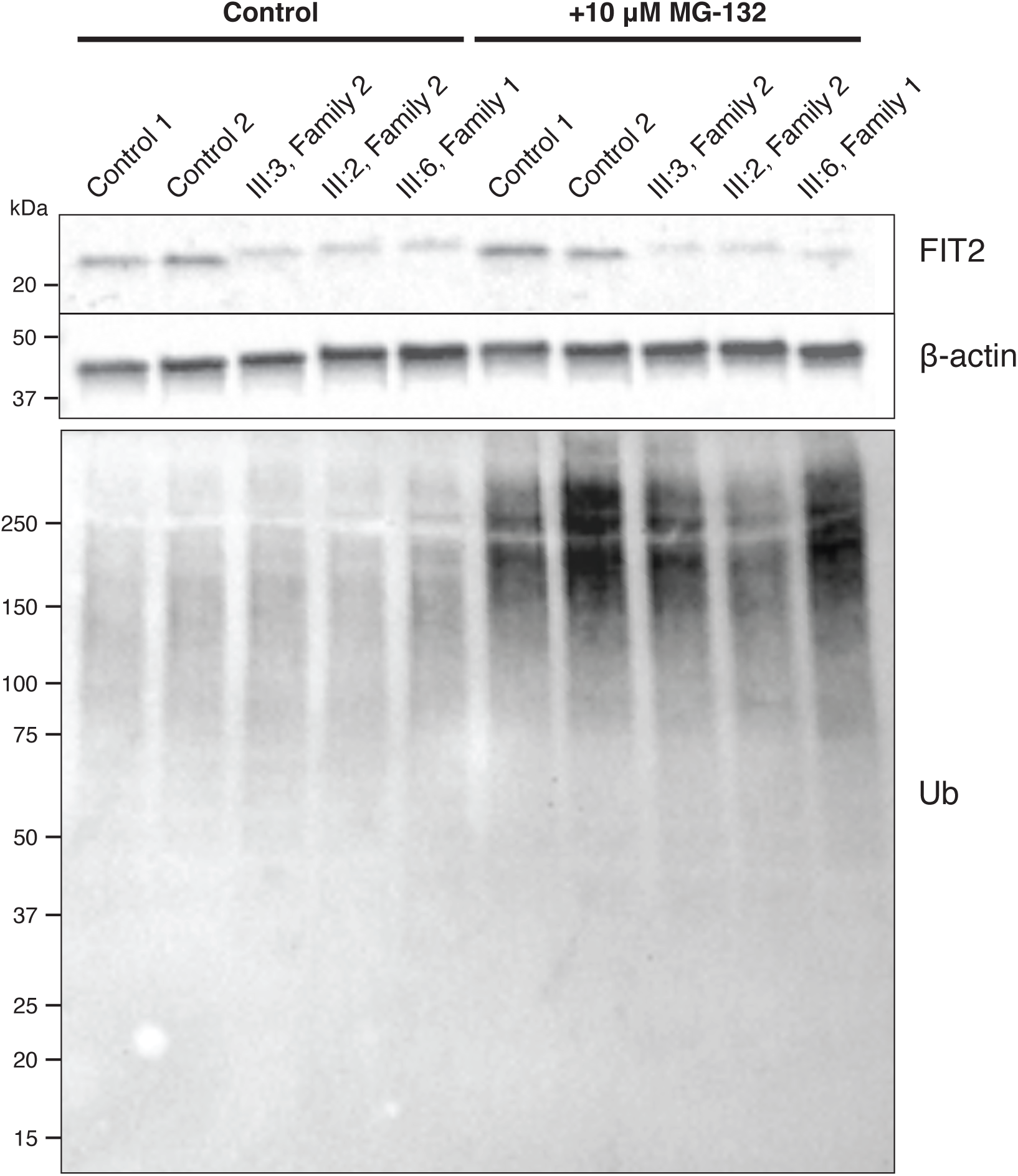
Immunoblot analysis of FIT2 in human fibroblasts with MG-132 treatment. Cells were treated for 24 hours with 10 μM MG-132. No significant changes were detected.

**Figure S6.**
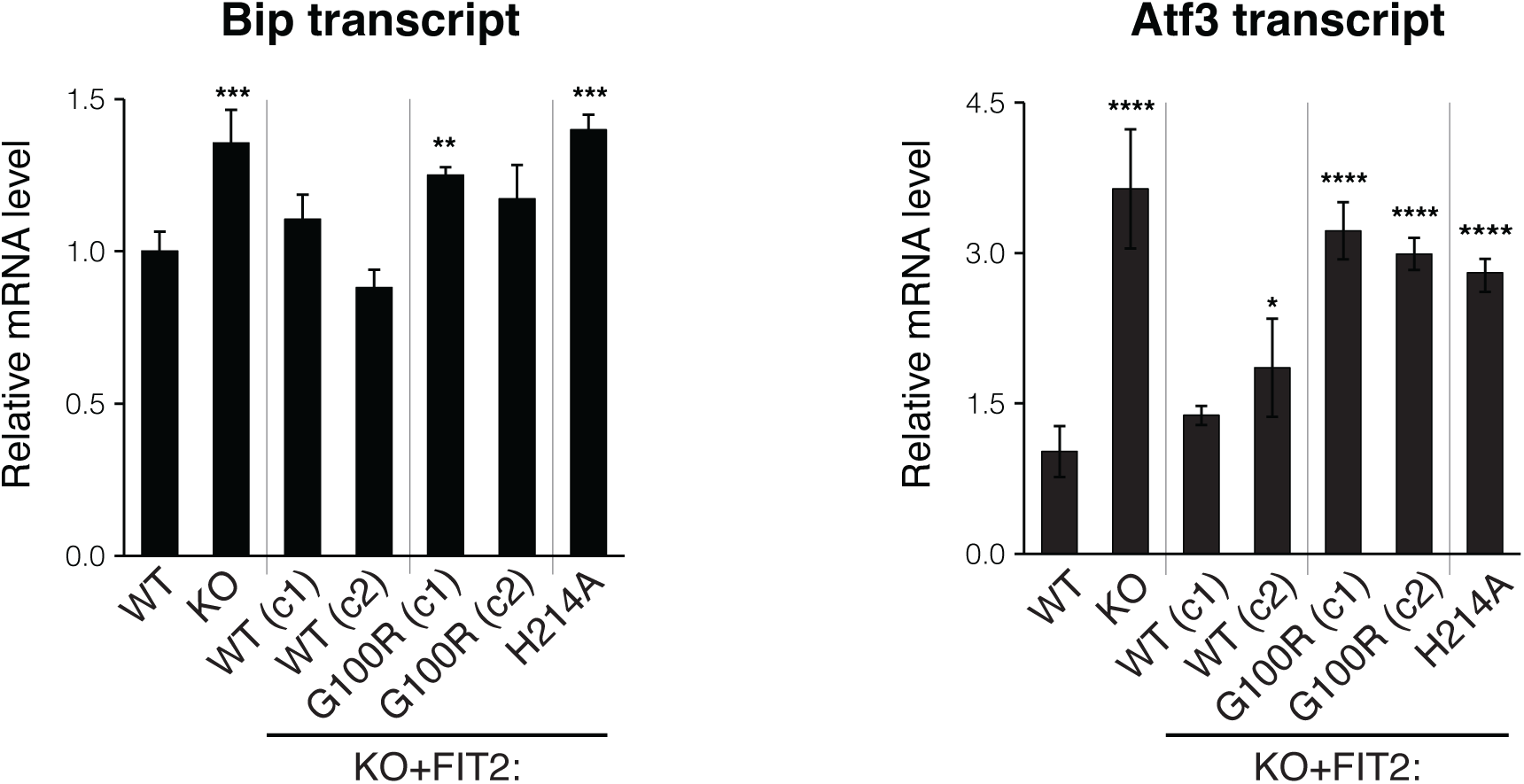
qRT-PCR analysis of Bip and Atf3 transcript levels. Both Bip and Atf3 transcript levels indicate that G100R-FIT2 does not completely rescue activation of the unfolded protein response exhibited by FIT2KO cells. Data represent mean ± SD, n = 3/cell line, *p <0.05, **p < 0.01, ***p < 0.001, and ****p < 0.0001, vs. WT, one-way ANOVA, followed by post-hoc Dunnett’s multiple comparisons.

**Figure S7.**
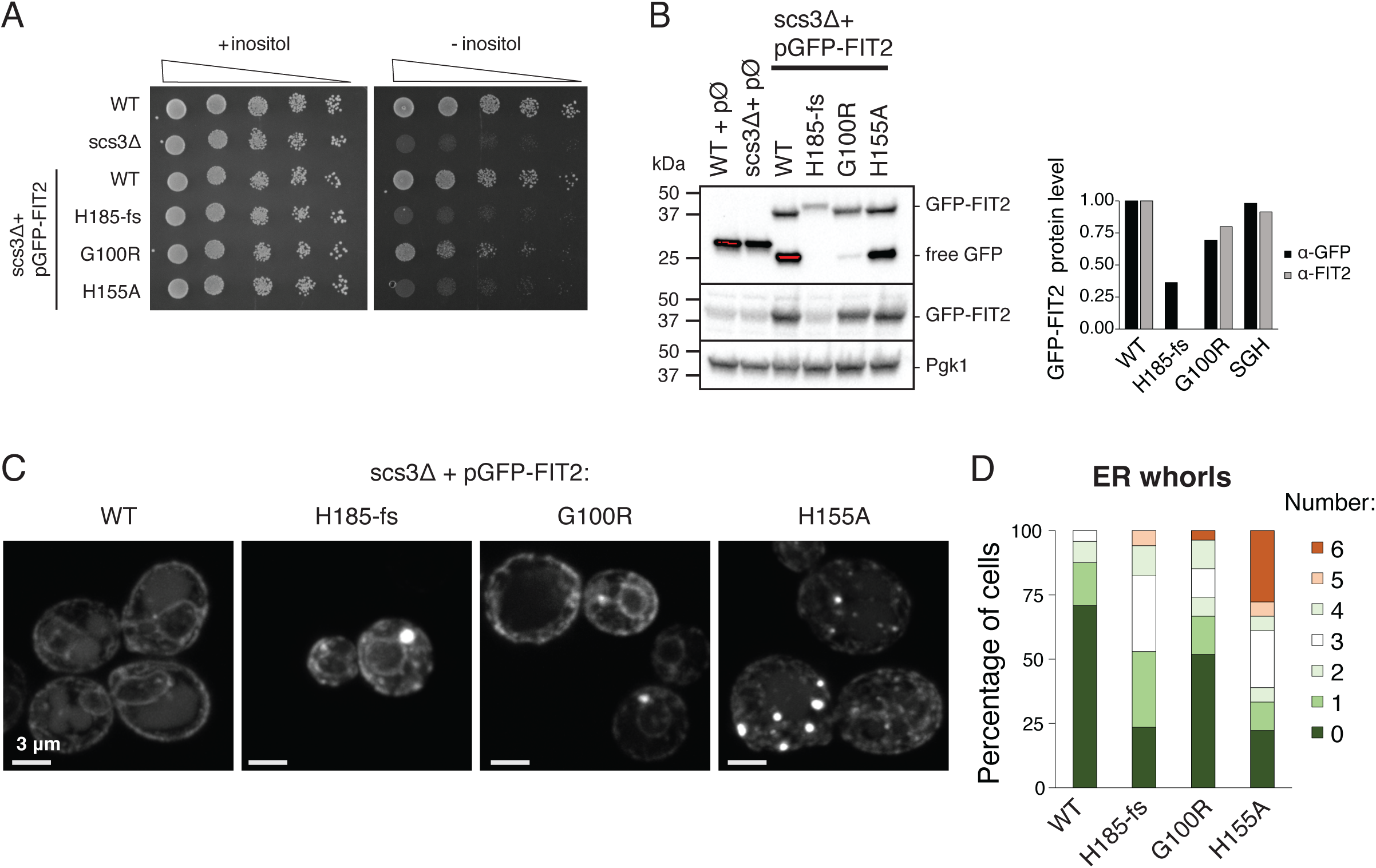
Molecular analysis of FIT2 mutations in yeast indicates that H105-fs-FIT2 is null and G100R-FIT2 is a hypomorph. **(A)** Drop assay indicates that H185-fs does not rescue, but G100R partially rescues, inositol auxotrophy in scs3Δ yeast. Cells were serially diluted and spotted on complete solid medium or inositol-depleted medium. **(B)** Immunoblot analysis of whole-cell lysates to determine GFP-FIT2 protein levels (normalized to PGK1) in scs3Δ cells expressing GFP-tagged FIT2 variants on a plasmid. Quantification of protein levels demonstrates near loss of GFP-FIT2-H185-fs and reduction of GFP-FIT2-G100R (FIT2 antibody does not recognize H185fs mutant). **(C)** Confocal imaging of scs3Δ cells expressing GFP-tagged FIT2 variants on a plasmid with exposure time adjusted to allow for visualization. GFP signal localizes to vacuole and ER in GFP-FIT2-WT and -H155A expression cells but ER signal in H185-fs and G100R cells. **(D)** Quantification of ER whorl number based on GFP-FIT2 signal from 20-30 cells per genotype.

